# Policy priorities and operational planning for socially determined diseases in Brazil: a nationwide analysis of municipal health plans

**DOI:** 10.64898/2026.06.25.26356624

**Authors:** Davi Rios do Nascimento, Gabriel de Jesus Gama dos Santos, Vinícius Pereira Lopes da Silva, Theo Oliveira Pieruccini Brisotto, Nayane Alves Cordeiro, Derlany José da Silva Belfort, Laura Moreira de Carvalho Soares, Paula Esbaltar de Oliveira, Sarah Fonseca Serpa, Carla Pacheco Teixeira, Diana Paola Gutierrez Diaz de Azevedo, Alessandra Xavier Bueno, Patrícia Rodrigues Sanine, Michael Ferreira Machado, Carmem Emmanuely Leitão Araújo, Thais Silva Matos, Rodrigo Feliciano do Carmo, Carlos Dornels Freire de Souza

**Affiliations:** Federal University of the São Francisco Valley. Petrolina, Pernambuco, Brazil; Oswaldo Cruz Foundation (Fiocruz), Rio de Janeiro, Rio de Janeiro, Brazil; Brazilian Ministry of Health. Brasília, Federal District, Brazil; São Paulo State University. São Paulo, SP, Brazil; Federal University of Alagoas. Maceió, Alagoas, Brazil; Federal University of Ceará. Fortaleza, Ceará, Brazil; University of Pernambuco. Petrolina, Pernambuco, Brazil

**Keywords:** Socially Determined Diseases, Health Planning, Institutional Capacity, Health Policy, Unified Health System

## Abstract

**Background:** Socially Determined Diseases (SDDs) remain closely associated with health inequities and require coordinated responses from health systems, even in countries with universal healthcare coverage. In Brazil, the Brasil Saudável Program (PBS) identified priority diseases and municipalities for elimination efforts, placing Municipal Health Plans (MHPs) at the centre of governance and policy implementation.

**Objective:** To analyse how and to what extent diseases prioritised by the PBS are incorporated into MHPs and translated into programmatic actions, considering municipal institutional capacity and service delivery organisation.

**Methods:** This descriptive documentary study analysed the MHPs of 175 municipalities prioritised by the PBS for the 2022–2025 planning cycle. Structured assessment instruments were developed based on the Program’s National Guidelines to evaluate planning attributes, alignment with national priorities, and the operationalisation of disease-specific actions. Descriptive analyses were performed using absolute and relative frequencies. Pearson’s chi-square test was used to assess associations between key planning attributes and regional differences.

**Results:** Although SDD-related goals were included in most plans, only 29.7% (n = 52) presented fully defined disease-specific targets, and 25.7% (n = 45) provided complete epidemiological information for all priority diseases. Dedicated funding was identified in 11.4% (n = 20) of municipalities, while fully structured awareness campaigns were described in only 4.0% (n = 7). Municipalities that incorporated epidemiological information into their plans were significantly more likely to describe structured prevention and control programmes than those without such information (76.7% versus 30.4%; χ^2^ = 29.8; p < 0.001). Regional differences were observed regarding the provision of disease-specific funding (χ^2^ = 12.0; p = 0.018), although planning profiles remained broadly similar across regions.

**Conclusions:** SDDs prioritised by the PBS are widely incorporated into MHPs at a declaratory level; however, their translation into structured programmatic actions remains limited. Weaknesses in financing, epidemiological intelligence, and operational planning constrain the implementation of national priorities. Strengthening municipal institutional capacity is essential to improve the operationalisation of health policies aimed at diseases shaped by social inequalities.

## Background

The social determinants of health perspective highlight the structural and systemic inequalities that shape patterns of illness, access to care, and health outcomes across populations (1). Within this broader analytical framework, the term Socially Determined Diseases (SDDs) has been used to designate a group of illnesses that disproportionately affect populations living in poverty and whose distribution reflects historical and structural social inequalities (2,3). Many of these conditions are still predominantly referred to in the international literature as Neglected Tropical Diseases (NTDs), defined as a set of diseases that, despite their profound global impact on morbidity, have historically received limited investment in research, development, and control efforts (4,5).

These conditions pose major challenges to global health, exerting a substantial burden even in countries with universal health systems. They affect more than one billion people worldwide and account for approximately 22 million disability-adjusted life years (DALYs) lost annually (4,6). In Brazil, an estimated 8,000 deaths per year are attributable to these diseases (7,8).

The magnitude of these diseases has driven global health responses, including the roadmap *“Ending the neglect to attain the Sustainable Development Goals: A road map for neglected tropical diseases 2021–2030”*, launched by the World Health Organization (WHO) in 2020. This initiative sets targets for the control, prevention, and elimination of these diseases by 2030, with emphasis on strengthening health systems, fostering international cooperation, expanding access to treatments, ensuring adequate financing, and promoting innovation (9).

In Brazil, the institutional response to the persistence of tuberculosis and other diseases associated with social vulnerability gained prominence from 2023 onward, with the establishment of the Interministerial Committee for the Elimination of Tuberculosis and Other Socially Determined Diseases (*Comitê Interministerial para a Eliminação da Tuberculose e de Outras Doenças Determinadas Socialmente* [CIEDS], in Portuguese). The committee was created to coordinate intersectoral actions to eliminate these diseases as public health problems by 2030 (10). To advance this agenda, the *Brasil Saudável – Unir para Cuidar Program* (PBS) was launched in 2024 with the objective of addressing socio-environmental determinants that disproportionately affect populations experiencing greater social vulnerability (11).

In its guidelines, the PBS identified 11 diseases: Chagas disease, schistosomiasis, lymphatic filariasis, soil-transmitted helminthiases, malaria, onchocerciasis, trachoma, tuberculosis, HIV/AIDS, viral hepatitis, and leprosy, and five vertically transmitted infections: Chagas disease, hepatitis B, HIV, HTLV infection, and syphilis, as being related to social conditions. These were collectively designated as SDD, and specific targets were established for their elimination. Additionally, 175 Brazilian municipalities with a high burden of two or more of these diseases were prioritised for planning and implementing strategies to achieve these goals (2).

While many of these conditions are traditionally discussed in the international literature under the umbrella of NTDs, the category of SDDs used in the Brazilian policy context is broader and includes conditions whose persistence is strongly shaped by social vulnerability, territorial inequalities, and barriers to access within the health system. The epidemiology of SDDs is closely linked to socioeconomic and socio-environmental conditions, necessitating intersectoral public policies focused on health promotion, prevention, and reducing structural inequalities (12). In this context, health planning plays a central role by enabling both the anticipation of emerging problems and the response to those already established. It guides the definition of priorities, goals, and strategies based on local realities, while promoting the rational allocation of resources and the coordination of health actions (13,14).

Within this framework, the Municipal Health Plan (MHP; *Plano Municipal de Saúde* [PMS], in Portuguese) serves as the primary strategic planning instrument for municipal management in the Unified Health System (*Sistema Único de Saúde* [SUS], in Portuguese). Prepared every four years as a mandatory requirement, it is responsible for organising and aligning health actions and services within the territory. The plan serves as an analytical and managerial tool that integrates social determinants, inequalities, and institutional capacities, while linking planning to budgeting and reaffirming the commitment to improving the population’s living conditions (15).

Accordingly, the analysis of MHPs may also serve as a proxy for local governmental capacity, as they reveal how health problems are identified, prioritised, and translated into programmatic actions within specific territorial and institutional contexts (16). In this context, the objective of this study was to analyse how and to what extent diseases prioritised by the PBS were incorporated into the MHPs of priority Brazilian municipalities and translated into programmatic actions. In this study, the SDD group extends beyond classical NTDs and is used as an analytical category aligned with national policy frameworks.

## Methods

### Study Design and Data Sources

This is a descriptive documentary study based on a comprehensive analysis of the MHPs of municipalities prioritised by the PBS. The MHPs for the 2022–2025 cycle, approved by the respective Municipal Health Councils – the SUS social oversight body – and made publicly available through transparency portals, were included in the analysis. When multiple versions were identified, the most recent version was selected.

It is important to note that the MHPs analysed were approved prior to the implementation of the PBS. Therefore, this study does not assess its outcomes or effects, but rather examines the extent to which the municipal planning context had already recognised and incorporated these diseases into local health plans. Furthermore, the PBS guidelines are embedded in other national policies and aligned with the Sustainable Development Goals and the SUS’s Organic Health Laws. These guidelines include: (1) addressing hunger and poverty; (2) reducing inequalities and expanding access; (3) strengthening communication; (4) promoting science, technology, and innovation; and (5) expanding infrastructure actions (2).

### Area of study

The unit of analysis in this study consisted of the MHPs of municipalities prioritised by the PBS. Of the 175 municipalities included in the analysis, the highest concentration was observed in the Southeast (29.71%; n = 52) and North (26.29%; n = 46) regions, followed by the Northeast (19.43%; n = 34), South (17.71%; n = 31), and Central-West (6.86%; n = 12) (2). Together, these municipalities account for approximately 93.7 million inhabitants (17), with marked variation in population size, ranging from small municipalities with limited administrative structure to large urban centres with greater technical and institutional capacity for health planning (18). For descriptive purposes, municipalities were classified according to population size as small (<50,000 inhabitants), medium (50,000–99,999 inhabitants), large (100,000–499,999 inhabitants), and very large (≥500,000 inhabitants). Population estimates and classifications are presented in Supplementary Material 3 (19).

**Fig. 1.**
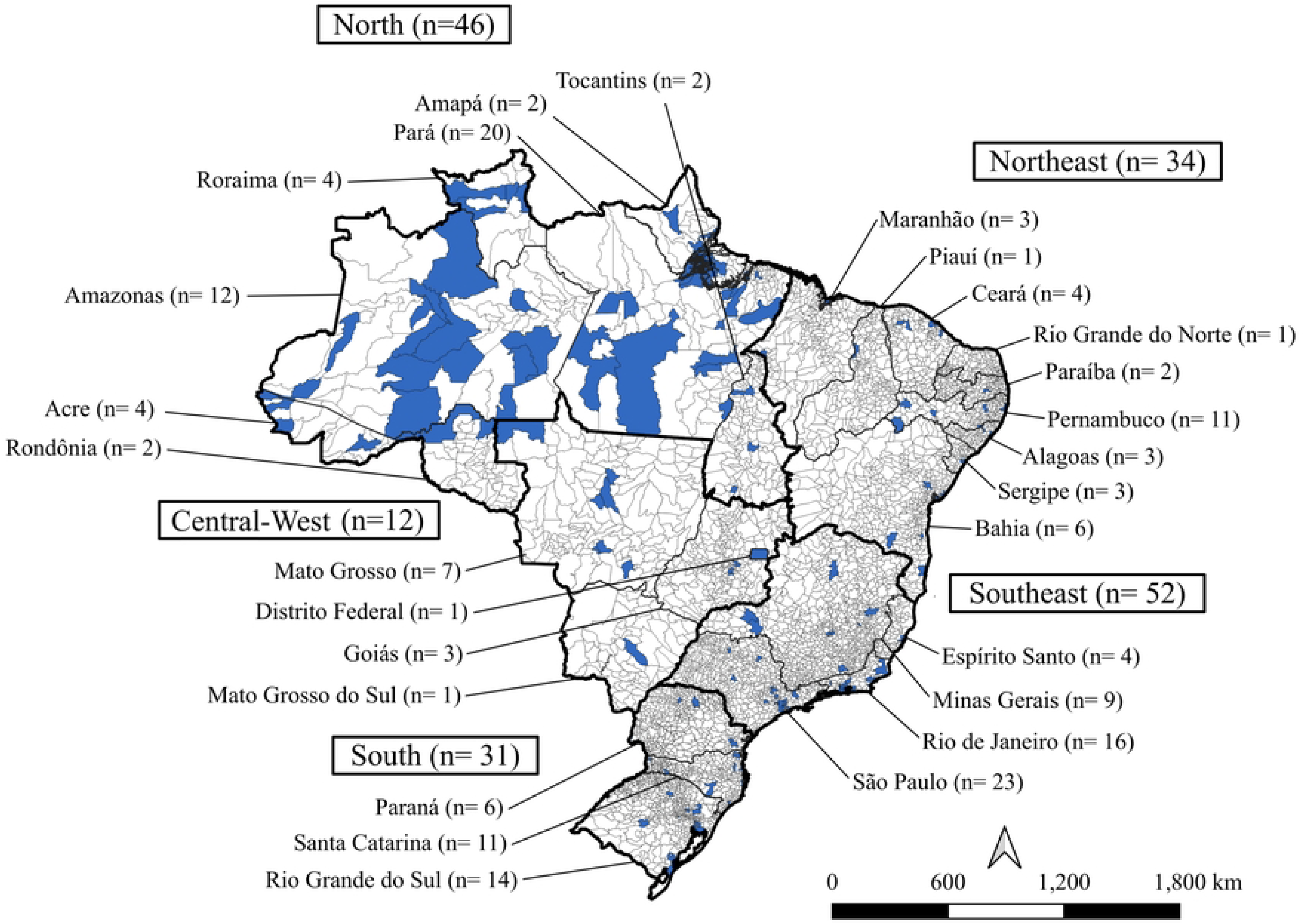
Priority municipalities of the Brasil Saudável – Unir para Cuidar Program by region. Source: National Guidelines of the PBS (2); authors’ elaboration.

### Data source

This study was based on the documentary analysis of publicly available secondary data, with the primary data source being the *DigiSUS Gestor – Módulo Planejamento* system (DGMP; https://digisusgmp.saude.gov.br/), the official platform of the Ministry of Health for registering health planning instruments. Data extraction was primarily conducted through this repository and, when necessary, supplemented by consultation of transparency portals and official municipal government websites, particularly in cases of technical inaccessibility or the absence of the document in the federal system. To ensure greater consistency in the search process, predefined descriptors were used, combining the terms “*Plano Municipal de Saúde*”, “PMS 2022–2025”, and “planning instruments.”

When the Municipal Health Plan for the 2022–2025 cycle was not publicly available either in the DGMP or through official municipal channels, the municipality remained in the sample and received a score of zero across all evaluated domains. This decision was based on the premise that the lack of public access to the planning instrument compromises transparency, accountability, and the ability to conduct technical assessment. Nevertheless, this approach should be interpreted with caution, as the lack of available documents does not necessarily imply the absence of planning activities. The municipalities of Maringá (Paraná) and Paulínia (São Paulo) met these criteria.

The consistency of the information was ensured by using only documents from official sources, which are typically submitted to the Municipal Health Councils for review and approval. When records were incomplete, dispersed, or fragmented, the analysis was limited to the content effectively available. Thematic domains that were entirely absent were assigned a score of zero, in accordance with the evaluation framework’s criteria, to preserve comparability across the municipalities analysed. This approach may overestimate deficiencies in planning capacity by conflating the absence of documentation with the absence of action. Data collection and systematisation were conducted between February and November 2025.

In this study, municipal institutional capacity was assessed indirectly through the content of the MHPs, considering the presence of an epidemiological situational analysis, explicit targets, funding allocation, intersectoral partnerships, and descriptions of operational actions related to priority diseases.

### Procedures and Steps of the Study

The study was organised into two main stages, through which documentary information was converted into analytical indicators of planning quality:

#### Stage 1 – Document Analysis and Scaling

The initial stage consisted of a systematic analysis of each Municipal Health Plan (MHP), using two forms developed by the authors, based on the information contained in the National Guidelines of the PBS (2). The questionnaires are presented in **Supplementary Materials 1 and 2**.

The questionnaire assessing the alignment of the MHPs with the National Guidelines of the PBS categorised responses as “Yes,” “Yes, for all,” “No,” and “Partially.” The category “Yes” was assigned when the assessed item was fully described in the MHPs for at least one of the diseases defined as priorities in that municipality, whereas “Yes, for all” was used when the evaluated content was completely and explicitly described for all priority diseases applicable to that municipality.

The category “Partially” was assigned based on a formal judgment criterion in cases where the item was explicitly mentioned in the document but presented insufficient technical detail or did not include the minimum content required for its operationalisation. This distinction enabled an objective assessment of both the presence and the extent of alignment between the MHPs and the national guidelines for the elimination of SDD.

In the questionnaire analysing the MHPs regarding SDD/Infections, municipalities previously identified as a priority for a given disease were assessed using specific items that addressed the presence of disease-related targets in their MHPs. Responses were classified as “Yes” or “No,” and the number of times the disease was mentioned in the MHPs was also recorded.

#### Stage 2 – Data Processing

A descriptive analysis of the responses was conducted using absolute and relative frequencies. This stage was essential for identifying the distribution of the variables across the five Brazilian regions, and for characterising the extent of programmatic incorporation of priority diseases.

Institutional capacity was indirectly inferred from planning attributes, including: (i) planning completeness, (ii) availability of epidemiological data, (iii) financial allocation, and (iv) operationalisation of actions.

For analytical purposes, PBS-prioritised diseases were grouped according to the degree of institutionalisation of their corresponding public health responses within the Brazilian Unified Health System (SUS). Diseases were considered to have more institutionalised responses when they were supported by long-standing national programmes, dedicated surveillance systems, established clinical guidelines, regular financing mechanisms, and consolidated service delivery structures. This group included tuberculosis, leprosy, HIV/AIDS, viral hepatitis, congenital syphilis, and malaria. Although malaria is geographically concentrated in specific endemic areas, it was classified as having a more institutionalised response within the SUS due to the existence of long-standing surveillance systems, dedicated financing mechanisms, established clinical protocols, and a consolidated national control programme.

Diseases were considered to have less institutionalised responses when characterised by less consolidated policy and service arrangements within the SUS, limited service organisation, lower visibility in routine health planning, or greater dependence on intersectoral actions. This group included HTLV, Chagas disease, schistosomiasis, lymphatic filariasis, trachoma, onchocerciasis, and soil-transmitted helminth infections.

Additional inferential analyses were performed to explore associations between key planning attributes. Pearson’s chi-square test was used to assess associations between the presence of disease-specific funding and the definition of targets, as well as between the availability of epidemiological data and the operationalisation of disease-specific programmes. Chi-square tests were also used to compare selected planning attributes across Brazilian macro-regions. Statistical significance was set at p < 0.05.

### Data Extraction Quality Control

To ensure the reliability of the information and the rigour of the analysis, data extraction followed a quality-control protocol organised into two stages. In the first stage, a double independent extraction was carried out on a random sample corresponding to 10% of the total MHPs evaluated (n = 18), with any discrepancies resolved through technical review and consensus among the evaluators. In the second stage, qualitative responses were validated through a new systematic review of justifications for items classified as “Partially,” aiming to standardise judgment criteria and ensure appropriate categorisation of the identified gaps.

No formal statistical metric was employed to estimate inter-rater agreement. Nevertheless, the procedures adopted aimed to minimise potential classification bias and ensure consistency and standardisation of analytical decisions.

### Ethical aspects

The study was based exclusively on secondary data obtained from publicly accessible information systems, in which individual identification is not possible. Therefore, ethical review or approval by a research ethics committee was not required.

## Results

Across the 175 priority municipalities analysed, substantial variability was observed in the extent to which the PBS guidelines were incorporated into the MHPs. The guideline related to reducing inequalities and expanding access to health services was the most frequently addressed, appearing in 89.1% (n = 156) of the plans. This was followed by guidelines aimed at expanding infrastructure actions, identified in 61.7% (n = 108) of the municipalities, and those focused on addressing hunger and poverty, as well as strengthening health communication, both present in 49.1% (n = 86). The guideline on promoting science, technology, and innovation had the lowest frequency, appearing in 34.9% (n = 61) of the MHPs. In 3.4% (n = 6) of the municipalities, none of the Program’s guidelines was identified in the planning instruments (**Table 1**).

**Table 1.** Guidelines of the PBS included in the MHPs, Brazil 2022-2025.

| Guidelines | n | % |
| --- | --- | --- |
| Guideline 1 – Hunger and poverty reduction | 86 | 49.1 |
| Guideline 2 – Reduction of inequalities and expansion of health access | 156 | 89.1 |
| Guideline 3 – Health communication enhancement | 86 | 49.1 |
| Guideline 4 – Promotion of science, technology, and innovation | 61 | 34.9 |
| Guideline 5 – Expansion of infrastructure actions | 108 | 61.7 |
| No guidelines identified | 6 | 3.4 |

Source: Authors' elaboration.

The assessment of the overall capacity of municipal planning to address SDD revealed a high frequency of reported intersectoral partnerships, identified in 78.9% (n = 138) of municipalities. The inclusion of general targets to reduce or eliminate these diseases was present in most MHPs, with 86.3% (n = 151) of the municipalities analysed. In contrast, the provision of specific funding for SDD-related actions was uncommon, identified in only 11.4% (n = 20) of the plans, whereas 88.6% (n = 155) did not include any budget allocation for these actions (**Table 2**).

**Table 2.** Overall capacity of municipal planning to address SDD according to the MHPs, Brazil 2022-2025.

|  | Answer | n | % |
| --- | --- | --- | --- |
| Existence of intersectoral partnerships for implementation of PBS guidelines. | Yes | 138 | 78.9 |
|  | No | 37 | 21.1 |
| Inclusion of specific targets for reducing or eliminating SDD in the MHPs. | Yes | 151 | 86.3 |
|  | No | 24 | 13.7 |
| Provision of specific funding for actions aimed at eliminating SDDs | Yes | 20 | 11.4 |
|  | No | 155 | 88.6 |

Source: Authors' elaboration.

Among the subset of municipalities in which diseases were defined as priorities by the PBS, heterogeneity was observed in how these priorities were translated into structured programmatic components. Specific targets for the reduction or elimination of priority diseases were fully described in 29.7% (n = 52) of the municipalities, while 58.9% (n = 103) presented partial target definitions and 11.4% (n = 20) did not include any specific targets. Likewise, the availability of updated epidemiological data in the MHPs was limited, with only 25.7% (n = 45) of the municipalities presenting complete information for all priority diseases, whereas 48.0% (n = 84) had partial data and 26.3% (n = 46) did not present updated epidemiological data (**Table 3)**.

**Table 3.** Programmatic translation of diseases defined as priorities by the PBS into the MHPs of priority municipalities, Brazil 2022-2025.

|  | Answer | n | % |
| --- | --- | --- | --- |
| Specific targets for priority diseases defined by the PBS | Yes, for all | 52 | 29.7 |
|  | Partially | 103 | 58.9 |
|  | No | 20 | 11.4 |
| Availability of up-to-date epidemiological data on priority diseases in the MHPs. | Yes, for all | 45 | 25.7 |
|  | Partially | 84 | 48.0 |
|  | No | 46 | 26.3 |
| Description of specific programs for the prevention and control of priority diseases. | Yes, for all | 37 | 21.2 |
|  | Partially | 76 | 43.4 |
|  | No | 62 | 35.4 |
| Description of awareness campaigns about priority diseases. | Yes, for all | 7 | 4.0 |
|  | Partially | 73 | 41.7 |
|  | No | 95 | 54.3 |
| Description of difficulties in implementing actions related to priority diseases. | Yes, for all | 14 | 8.0 |
|  | Partially | 48 | 27.4 |
|  | No | 113 | 64.6 |
| Provision for training health professionals to implement the targets | Yes | 162 | 92.6 |
|  | No | 13 | 7.4 |

Source: Authors' elaboration.

Regarding the operationalisation of actions, a full description of specific prevention and control programs was present in 21.1% (n = 37) of municipalities, while 43.4% (n = 76) provided a partial description, and 35.4% (n = 62) did not report the existence of structured programs. Awareness campaigns were even less frequent, with a full description in only 4.0% (n = 7) of municipalities, a partial description in 41.7% (n = 73), and no mention in 54.3% (n = 95). The explicit reporting of implementation difficulties was infrequent, being partially described in 27.4% (n = 48) and absent in 64.6% (n = 113). In contrast, provision for the training of health professionals was observed in 92.6% (n = 162) of the municipalities analysed (**Table 3**).

Additional inferential analyses identified a significant association between the availability of epidemiological data and the operationalisation of disease-specific actions. Municipalities that presented complete or partial epidemiological information on priority diseases were more likely to describe structured prevention and control programmes than municipalities without epidemiological data (76.7% versus 30.4%; χ^2^ = 29.8; p < 0.001) **(Table 4**).

**Table 4.**
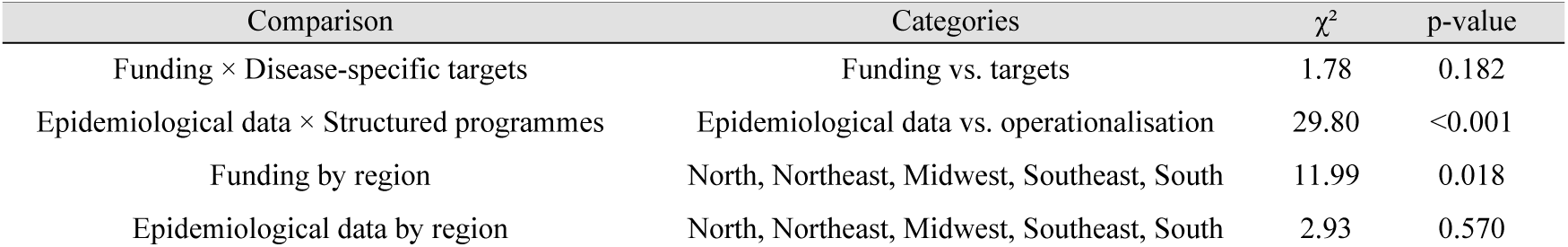
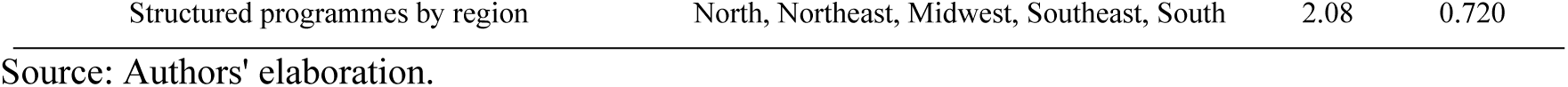
Inferential analysis of planning attributes and regional differences in Municipal Health Plans, Brazil, 2022–2025.

Regional differences were observed regarding the provision of specific funding for SDD-related actions (χ^2^ = 12.0; p = 0.018). The Northeast region showed the highest proportion of municipalities reporting dedicated funding (26.5%), whereas no municipality in the South region reported specific financial allocation for these actions. No significant association was observed between the presence of disease-specific funding and the definition of targets (χ^2^ = 1.78; p = 0.182). Likewise, no significant regional differences were identified regarding the availability of epidemiological data (χ^2^ = 2.93; p = 0.570) or the existence of disease-specific programmes (χ^2^ = 2.08; p = 0.720) (**Table 4)**.

Substantial heterogeneity was observed in the degree of programmatic incorporation across PBS-prioritised diseases. Diseases with more institutionalised policy responses within the SUS, such as tuberculosis, HIV/AIDS, viral hepatitis, congenital syphilis, and leprosy, generally presented higher levels of incorporation into municipal planning. In contrast, diseases with less institutionalised policy arrangements, particularly those dependent on intersectoral and socio-environmental interventions, showed limited operationalisation, even in municipalities where they had been defined as priorities by the Program (**Table 5**).

**Table 5.**
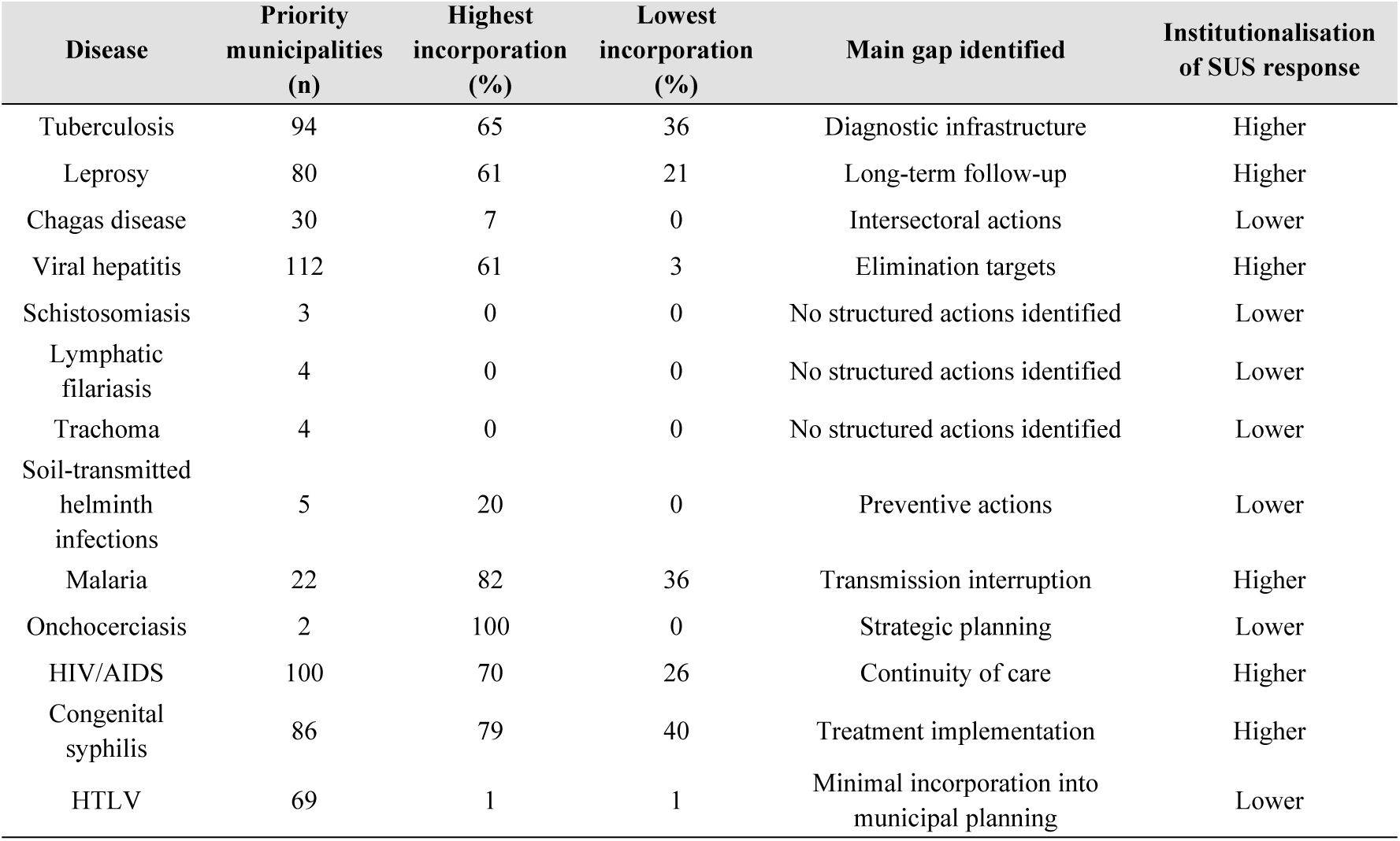
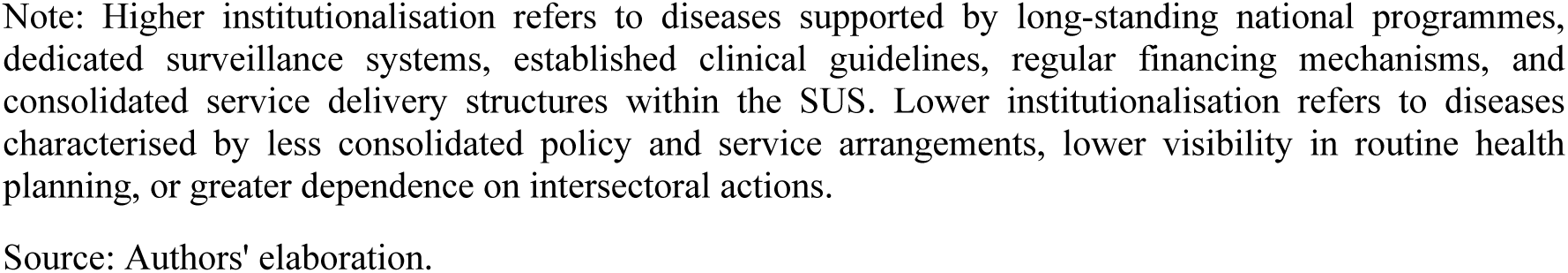
Institutionalisation of SUS responses and programmatic incorporation of PBS-prioritised diseases in Municipal Health Plans, Brazil, 2022–2025.

| Disease | Priority municipalities (n) | Highest incorporation (%) | Lowest incorporation (%) | Main gap identified | Institutionalisation of SUS response |
| --- | --- | --- | --- | --- | --- |
| Tuberculosis | 94 | 65 | 36 | Diagnostic infrastructure | Higher |
| Leprosy | 80 | 61 | 21 | Long-term follow-up | Higher |
| Chagas disease | 30 | 7 | 0 | Intersectoral actions | Lower |
| Viral hepatitis | 112 | 61 | 3 | Elimination targets | Higher |
| Schistosomiasis | 3 | 0 | 0 | No structured actions identified | Lower |
| Lymphatic filariasis | 4 | 0 | 0 | No structured actions identified | Lower |
| Trachoma | 4 | 0 | 0 | No structured actions identified | Lower |
| Soil-transmitted helminth infections | 5 | 20 | 0 | Preventive actions | Lower |
| Malaria | 22 | 82 | 36 | Transmission interruption | Higher |
| Onchocerciasis | 2 | 100 | 0 | Strategic planning | Lower |
| HIV/AIDS | 100 | 70 | 26 | Continuity of care | Higher |
| Congenital syphilis | 86 | 79 | 40 | Treatment implementation | Higher |
| HTLV | 69 | 1 | 1 | Minimal incorporation into municipal planning | Lower |

Note: Higher institutionalisation refers to diseases supported by long-standing national programmes, dedicated surveillance systems, established clinical guidelines, regular financing mechanisms, and consolidated service delivery structures within the SUS. Lower institutionalisation refers to diseases characterised by less consolidated policy and service arrangements, lower visibility in routine health planning, or greater dependence on intersectoral actions.
Source: Authors' elaboration.

The analysis of specific programmatic actions for each condition was conducted exclusively within the subset of priority municipalities in which the respective disease or infection had been designated a priority by the PBS, as listed in **Supplementary Material 3**.

The regional analysis revealed specific contrasts in the programmatic incorporation of diseases defined as priorities by the PBS, but these did not constitute qualitatively distinct regional profiles. Overall, conditions with longer-standing surveillance, financing, and care arrangements within the SUS showed a greater presence of targets and programmatic actions, whereas diseases strongly associated with complex socio-environmental determinants remained poorly operationalised, even in municipalities where they had previously been defined as priorities by the Program (**Supplementary Material 4**).

In the North region, greater structuring of malaria-targeted actions was observed, with the definition of targets and control strategies in a substantial proportion of municipalities where the condition was identified as a priority. In contrast, Chagas disease showed low operationalisation, with limited description of surveillance strategies and a lack of structured actions for vector control and housing improvement (**Supplementary Material 4**).

Overall, diseases with more consolidated trajectories within the Brazilian Unified Health System, such as tuberculosis, HIV/AIDS, and viral hepatitis, which benefit from longer-standing programs, clearer care pathways, and more stable financing and surveillance structures, were more readily incorporated into municipal planning. In contrast, diseases more dependent on intersectoral and territorially tailored responses remained less operationalised, even in municipalities where they had been prioritised. In the Northeast, leprosy stood out for its greater programmatic incorporation, particularly regarding active case finding and contact investigation, whereas conditions such as schistosomiasis and lymphatic filariasis remained largely absent from operational planning, even in municipalities where they had previously been defined as priorities. In the Southeast and South regions, diseases such as tuberculosis and HIV/AIDS showed greater programmatic incorporation, although relevant gaps persist in the regular provision of diagnostic testing and in ensuring continuous access to treatment (**Supplementary Material 4**).

## Discussion

This study analysed how, and to what extent, the response to SDD is incorporated into the MHPs of priority municipalities in Brazil and translated into programmatic actions. The results indicate that, although guidelines related to SDD are widely recognised in the MHPs of priority municipalities, their operationalisation remains limited, revealing structural weaknesses in municipal planning. General targets and declared partnerships predominated, in contrast to the limited provision of specific funding, the partial or absent definition of disease-specific targets, the limited availability of complete epidemiological data, and the insufficient detailing of operational strategies.

Within the field of health planning, these findings point to a persistent gap between federal norm-setting and local implementation capacity in decentralised systems. Although Municipal Health Plans frequently incorporate statements aligned with national priorities, translating these commitments into feasible strategies remains uneven. This pattern can be interpreted in light of the literature on policy capacity, implementation, and state capacities in subnational governments, which emphasises the interaction between analytical, managerial, political, and institutional dimensions, as well as the unequal bureaucratic, fiscal, and administrative conditions across Brazilian municipalities (20,21).

This pattern suggests the predominance of a declaratory mode of planning, driven more by formal compliance with normative requirements than by the effective operationalisation of actions. The limited presence of explicit funding, structured campaigns, and detailed implementation strategies indicates that priority-setting is often not accompanied by the managerial and institutional arrangements required for execution (22,23). This predominantly declarative nature of planning is corroborated by the low presence of concrete operational instruments, such as structured campaigns, which were fully described in only 4.0% of the analysed municipalities.

The inferential analyses reinforce this interpretation. Municipalities that incorporated epidemiological information into their health plans were significantly more likely to describe structured prevention and control programmes, suggesting that analytical capacity may be an important determinant of the operationalisation of local health actions.

Empirical evidence from Brazilian municipal contexts underscores the extent of the shortcomings observed in the structuring of MHPs. A study of MHPs from municipalities in the state of Minas Gerais found that only 10.0% of the documents presented a complete structure in accordance with national guidelines and only 16.7% contained fully defined guidelines, objectives, targets, and indicators, suggesting persistent limitations in the technical and operational capacity of municipal health planning (24). This interpretation is also consistent with the Brazilian literature on federalism and state capacity, which documents marked inequalities in bureaucracy, fiscal capacity, and administration across municipalities (18).

These shortcomings should also be interpreted in light of the substantial heterogeneity among Brazilian municipalities. As the literature on Brazilian federalism and state capacity has shown, subnational governments operate under highly unequal bureaucratic, fiscal, and socioeconomic conditions, and many municipalities still face precarious administrative structures. Such disparities directly affect their ability to consistently formulate, finance, implement, and monitor health plans (18).

Regional differences in the provision of disease-specific funding were also observed, with municipalities in the Northeast reporting dedicated financial allocation more frequently than those in other regions. This finding reinforces the existence of territorial heterogeneity in planning and resource allocation processes, suggesting that the incorporation of financial commitments to PBS-prioritised diseases may be influenced by regional governance arrangements and local administrative capacities (18).

These results further reinforce the relevance of the PBS as a national public policy by showing that its implementation occurs against a baseline of municipal planning characterised by significant operational gaps. As a baseline analysis, the study does not assess the Program’s outcomes or effects; rather, it identifies the existing capacities and limitations of municipal planning prior to the adoption of PBS guidelines, thereby providing an empirical reference point for future evaluations of policy implementation.

This diagnosis acquires greater relevance when considered in light of the persistence of uneven epidemiological patterns across the national territory. Epidemiological evidence indicates that, even within a universal health system, diseases strongly determined by socio-environmental conditions maintain persistent patterns of territorial concentration and inequality (25). Regarding tuberculosis, for example, space-time analyses have demonstrated high and stable spatial autocorrelation (global Moran’s I > 0.40), with approximately one quarter of cases concentrated in just over 15% of Brazilian municipalities, particularly in areas of pronounced social vulnerability (26).

Concurrently, Chagas disease remains a major public health problem in the country, with an estimated 1.1 to 4.6 million infected individuals and progression to chronic forms in approximately 30–40% of cases, reflecting longstanding limitations in surveillance, access to diagnosis, and the organisation of care (27). These findings are situated within a broader pattern in which countries with universal health systems concentrate a substantial share of the global burden of poverty-related diseases, accounting, according to analyses based on the Global Burden of Disease Study 2013, for approximately 60% of DALYs due to tuberculosis, 43–44% due to HIV/AIDS, and up to 78% due to Chagas disease (28).

In this context, the PBS should be understood not only as a coordination initiative, but also as a policy instrument with the potential to induce municipal capacity-building and reorienting local planning practices, especially for conditions whose elimination depends on sustained intersectoral and intergovernmental responses, as suggested by the literature on the challenges of schistosomiasis control in Brazil (29).

Its relevance lies precisely in confronting a baseline scenario marked by fragmented planning, weak operational detail, and limited integration between national priorities and local implementation. For the health system, these findings reinforce the need to strengthen technical support, earmarked financing, and intersectoral coordination mechanisms in order to reduce the gap between nationally defined priorities and their translation into consistent programmatic actions at the local level (30,31).

This study has limitations inherent to documentary analysis. MHPs express formal intentions, priorities, and planned actions, but do not allow direct inferences about implementation quality or program outcomes. In addition, the analysis focused on plans produced prior to the implementation of the PBS, which precludes any assessment of the Program’s direct effects. Finally, the absence of publicly available planning documents in a small number of municipalities may have affected the assessment of planning capacity, although this lack of transparency is itself relevant from a governance perspective. The assessment instruments were specifically developed for this study based on the National Guidelines of the Brasil Saudável Program and did not undergo formal psychometric validation. Although no formal agreement coefficient was calculated, future studies should incorporate inter-rater reliability measures.

Conversely, this same characteristic represents one of its main contributions, as it provides an analytical baseline of the municipal planning context that shapes the policy’s relevance. As an additional contribution, the study proposes a systematic framework for assessing the alignment between national guidelines and local planning instruments, with potential applicability to other contexts and to countries facing similar challenges in addressing diseases strongly shaped by social inequalities.

## Conclusion

This study showed that SDDs prioritised by the PBS are widely incorporated into Municipal Health Plans at a declaratory level; however, their translation into structured programmatic actions remains limited. General goals and references to national guidelines predominated, whereas disease-specific targets, dedicated funding, and operational instruments were frequently partial or absent. The availability of epidemiological information was significantly associated with the presence of structured prevention and control programmes, highlighting the importance of analytical capacity for translating planning priorities into operational actions. Regional differences were observed mainly in the intensity of incorporation rather than in qualitatively distinct planning profiles.

Future studies should examine how national policy induction influences municipal planning capacity over time and should explore, through qualitative and mixed-methods approaches, the institutional and territorial factors that shape the conversion of national priorities into concrete local action.

## Data Availability

All data underlying the findings of this study are publicly available. The Municipal Health Plans analyzed were obtained from the DigiSUS Gestor Planning Module (https://digisusgmp.saude.gov.br/) and, when unavailable through DigiSUS, from official municipal government transparency portals and websites. The list of municipalities analyzed, the analytical framework, and the evaluation instruments developed for this study are provided as Supporting Information files. Additional information regarding data extraction procedures is available from the corresponding author upon reasonable request.

## Declarations

### Ethics approval and consent to participate

This study used only secondary data obtained from publicly available information systems and official institutional documents, without access to identifiable individual data. Therefore, according to Brazilian regulations, ethics committee approval and informed consent were not required.

### Availability of data and materials

All data underlying the findings of this study are publicly available. The data analyzed were obtained from the DigiSUS Gestor – Planning Module (https://digisusgmp.saude.gov.br/) and official municipal transparency portals. All Municipal Health Plans analyzed are available through their respective institutional repositories. The analytical framework and evaluation instruments developed for this study are provided as Supporting Information. Additional information is available from the corresponding author upon reasonable request.

### Authors’ contributions

**Davi Rios do Nascimento (DRN):** Conceptualization, Methodology, Data curation, Formal analysis, Visualization, Writing - original draft, and Writing - review & editing.

**Gabriel de Jesus Gama dos Santos (GJGS):** Data curation, Formal analysis, and Writing - review & editing.

**Vinícius Pereira Lopes da Silva (VPLS):** Data curation, Formal analysis, and Writing - review & editing.

**Theo Oliveira Pieruccini Brisotto (TOPB):** Data curation, Formal analysis, and Writing - review & editing.

**Nayane Alves Cordeiro (NAC):** Data curation, Formal analysis, and Writing - review & editing.

**Derlany José da Silva Belfort (DJSB):** Data curation, Formal analysis, and Writing - review & editing.

**Laura Moreira de Carvalho Soares (LMCS):** Data curation, Formal analysis, and Writing - review & editing.

**Paula Esbaltar de Oliveira (PEO):** Data curation, Formal analysis, and Writing - review & editing.

**Sarah Fonseca Serpa (SFS):** Data curation, Formal analysis, and Writing - review & editing.

**Carla Pacheco Teixeira (CPT):** Writing - review & editing.

**Diana Paola Gutierrez Diaz de Azevedo (DPGDA):** Writing - review & editing.

**Alessandra Xavier Bueno (AXB):** Writing - review & editing.

**Patrícia Rodrigues Sanine (PRS):** Methodology, Writing - review & editing.

**Michael Ferreira Machado (MFM):** Validation, Writing - review & editing.

**Carmem Emmanuely Leitão Araújo (CELA):** Validation, Writing - review & editing.

**Thaís Silva Matos (TSM):** Methodology, Data curation, Formal analysis, Writing - review & editing, and Supervision.

**Rodrigo Feliciano do Carmo (RFC):** Methodology, Data curation, Formal analysis, Writing - review & editing, and Supervision.

**Carlos Dornels Freire de Souza (CDFS):** Conceptualization, Methodology, Supervision, Project administration, Validation and Writing - review & editing.

## Acknowledgements

Not applicable.

## AI declaration

During the preparation of this manuscript, generative artificial intelligence tools were used exclusively for language refinement and editorial assistance. All scientific content, interpretations, and conclusions were critically reviewed and validated by the authors, who assume full responsibility for the final version of the manuscript.

## Supplementary Material

**Supplementary Material 1-** Questionnaire for analysing Municipal Health Plans regarding the National Guidelines of the *Brasil Saudável – Unir para Cuidar* Program (.docx)

Description: Assessment instrument used to evaluate the alignment of Municipal Health Plans with the National Guidelines of the Brasil Saudável Program, including the identification of priority actions, intersectoral partnerships, targets, and funding provisions.

**Supplementary Material 2-** Questionnaire for analysing Municipal Health Plans regarding Socially Determined Diseases/Infections (.docx)

Description: Disease-specific assessment instrument used to evaluate the incorporation of PBS-prioritised diseases and infections into Municipal Health Plans, including targets, surveillance actions, prevention strategies, and service organisation components.

**Supplementary Material 3-** Priority municipalities by region in Brazil and their respective socially determined diseases/infections (.docx)

Description: List of the 175 municipalities prioritised by the Brasil Saudável Program, including region, population size classification, and the diseases and infections defined as priorities for each municipality.

**Supplementary Material 4-** Specific programmatic components for each disease, by priority disease and region (.docx)

Description: Detailed results of the documentary analysis showing the presence of disease-specific targets, surveillance activities, prevention and control actions, and other programmatic components according to priority disease and Brazilian macro-region.

